# Sex Differences in Management, Time to Intervention, and In-Hospital Mortality of Acute Myocardial Infarction and Non-Myocardial Infarction Related Cardiogenic Shock

**DOI:** 10.1101/2024.10.11.24315358

**Authors:** Anushka Desai, Rohan Rani, Anum Minhas, Faisal Rahman

**Affiliations:** Georgetown University School of Medicine; Division of Cardiology, Johns Hopkins University School of Medicine, Baltimore, Maryland, USA

**Author notes:** **Corresponding Author Informatio**n Anushka Desai, 3603 T Street NW Washington D.C., 20007. **Disclosures** None.

## Abstract

**Background:** Limited data are available on sex differences in the time to treatment of cardiogenic shock (CS) with and without acute myocardial infarction (AMI).

**Methods:** For this retrospective cohort study, we used nationally representative hospital survey data from the National Inpatient Sample (years 2016-2021) to assess sex differences in interventions, time to treatment (within versus after 24 hours of admission), and in-hospital mortality for AMI-CS and non-AMI-CS, adjusting for age, race, income, insurance, comorbidities, and prior cardiac interventions.

**Results:** We identified 1,052,360 weighted CS hospitalizations (60% non-AMI-CS; 40% AMI-CS). Women with CS had significantly lower rates of all interventions. For AMI-CS, women had a higher likelihood of in-hospital mortality after: revascularization (adjusted odds ratio (aOR) 1.15 [95% CI 1.09-1.22]), mechanical circulatory support (MCS) (1.15 [1.08-1.22]), right heart catheterization (RHC) (1.10 [1.02-1.19]) (all p<0.001). Similar trends were found for the non-AMI-CS group. Women with AMI-CS were less likely to receive early (within 24 hours of admission) revascularization (0.93 [0.89-0.96]), MCS (0.76 [0.73-0.80]), or RHC (0.89 [0.84-0.95]) than men; women with non-AMI-CS were less likely to receive early revascularization (0.78 [0.73-0.84]), IABP (0.85 [0.78-0.94]), pLVAD (0.88 [0.77-0.99]) or RHC (0.83 [0.79-0.88]) than men (all p<0.001). For both types of CS, in-hospital mortality was not significantly different between men and women receiving early ECMO, pLVAD, or PCI.

**Conclusions:** Sex disparities in the frequency of treatment of CS persist on a national scale, with women being more likely to die following treatment and less likely to receive early treatment. However, when comparing patients who received early treatment, in-hospital mortality does not differ significantly when men and women are treated equally within 24 hours of admission. Early intervention if clinically indicated could mitigate sex-based differences in CS outcomes and should be made a priority in the management of CS.

**Clinical Perspective:** What is new?

- Women have increased in-hospital mortality with cardiogenic shock and have lower odds of early intervention (within 24 hours of admission) compared to men.
- When men and women with cardiogenic shock are treated equally in terms of early intervention within 24 hours of admission, sex differences in in-hospital mortality disappear.

What are the clinical implications?

- Sex-based disparities are present in the utilization of treatment and time to intervention for patients with cardiogenic shock.
- Early recognition and intervention for cardiogenic shock among women should be considered to improve clinical outcomes.

## Introduction

Cardiogenic shock (CS) is defined as a state of decreased circulation causing hypoxia and end-organ hypoperfusion. It is frequently precipitated by acute myocardial infarction (AMI).^1^ Despite advancements in medical interventions over the past 20 years, CS remains associated with high mortality ranging between 30-50%.^2-5^ There exist marked sex disparities in mortality associated with CS in the United States.^6-8^ Current literature suggests insufficient recognition of symptoms in women, faltering guideline-directed medical therapy during the first 24 hours, and inadequate utilization of mechanical circulatory support (MCS) devices as underlying reasons for the observed mortality difference.^9^

In all patients, rapid deterioration following CS necessitates timely intervention to improve clinical outcomes.^10,11^ Recently, the DanGer Shock trial has driven the point of early aggressive treatment of AMI-CS, finding that prompt routine use of percutaneous left ventricular assist devices (pLVAD) decreases the risk of all-cause mortality.^12^ This is especially pertinent when considering a recent cohort study that found women to be less likely to receive pLVAD when hospitalized for CS.^13^ Given that earlier intervention can result in improved clinical outcomes, we aimed to investigate whether differences in timely intervention were associated with higher mortality among women compared to men.

## Methods

### Study Data

Data was obtained from the Healthcare Cost and Utilization Project (HCUP) National Inpatient Sample (NIS) database, which is sponsored by the Agency of Healthcare Research and Quality. The NIS is the largest publicly available all-payer administrative database in the United States. It contains hospitalization data from a stratified 20% sample of over 1,000 hospitals across the nation participating in the HCUP, and, when weighted, can be used to estimate nationwide trends and incidence.

### Study Population & Outcomes

The database was queried for adult patients (≥18 years) admitted with CS from 2016 to 2021, using the International Classification of Disease 10th Revision Clinical Modification Codes of R57.0. We categorized these hospitalizations by the presence of AMI on admission into two cohorts: non-AMI CS and AMI-CS. Hospitalizations missing mortality or sex data were excluded. Study flowchart is included in **Figure S1**.

Baseline demographics, comorbidities, and prior interventions were identified with ICD-10-CM codes. These characteristics included census-defined age group, race/ethnicity, quartile of median household income, insurance, hospital type, hospital teaching status, obesity, dyslipidemia, diabetes mellitus, hypertension, tobacco use, peripheral vascular disease, chronic heart failure, chronic kidney disease, chronic liver disease, valvular heart disease, coronary artery disease, stroke, prior percutaneous intervention (PCI), and prior coronary artery bypass graft (CABG).

The procedural outcomes analyzed were revascularization (Percutaneous Coronary Intervention [PCI] and Coronary Artery Bypass Graft [CABG]), MCS (Intra-aortic Balloon Pump [IABP], Percutaneous Left Ventricular Assist Device [pLVAD], Extracorporeal Membrane Oxygenation [ECMO]), RHC, and advanced heart failure therapies (LVAD and heart transplant). The clinical outcomes studied included in-hospital mortality, use of invasive mechanical ventilation, major bleeding, acute kidney injury, and stroke. **Table S1** includes a complete list of all ICD-10 diagnosis and procedure codes used.

### Statistical Analysis

All data represent weighted national estimates, which were done using HOSP_NIS as a clustering variable and NIS_STRATUM to account for different strata, as recommended by the AHRQ methods.^14^ Differences in baseline data, procedural outcomes, and clinical outcomes between the type of CS and sex were compared using Pearson’s χ^2^ test for categorical variables and the Mann-Whitney test for continuous variables.

Multivariable logistic regressions that accounted for survey weighting were used to assess sex differences in use of intervention at any point during hospitalization, early treatment (within 24 hours of admission), and adverse clinical outcomes for both AMI and non-AMI CS, adjusting for all baseline demographics, comorbidities, and prior interventions. Further subgroup analyses were performed to assess sex differences in in-hospital mortality associated with receiving early versus late intervention.

Statistical significance was defined with a type I error of < 0.05. All analyses were performed using STATA Statistical Software version 18.0.

## Results

### Baseline Characteristics

Our study included 1,052,360 weighted hospitalizations with CS between 2016 and 2021. As seen in **Table 1**, both non-AMI-CS and AMI-CS had a higher incidence in men compared to women (non-AMI-CS 61.8% vs 38.1%; AMI-CS 63.6% vs 36.3%; p<0.001). Women were more likely to be older with CS. Women with CS, compared to men, had higher frequencies of obesity, valvular heart disease, and stroke, and lower frequencies of coronary artery disease and prior PCI or CABG (p<0.001).

**Table 1.**
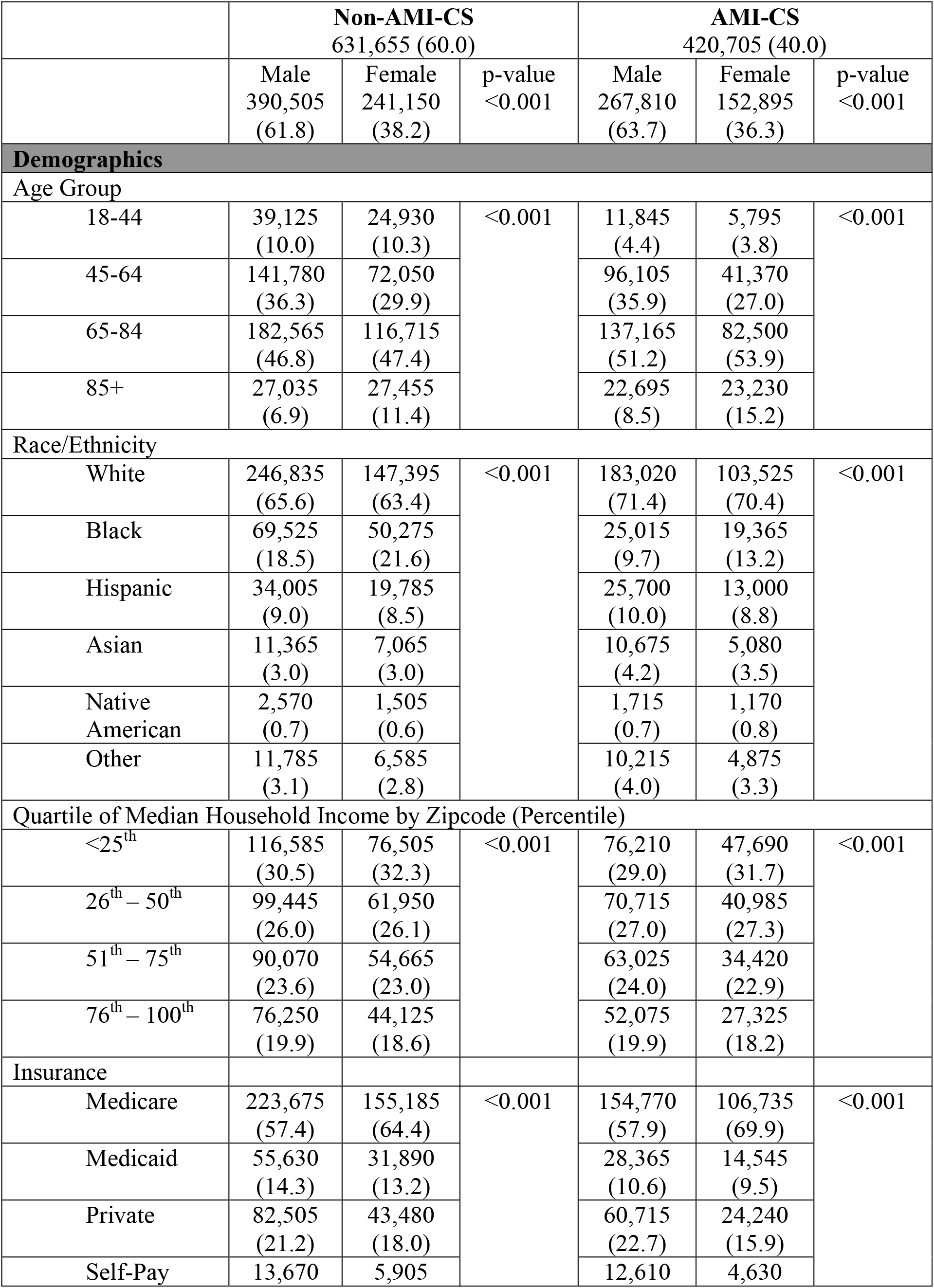
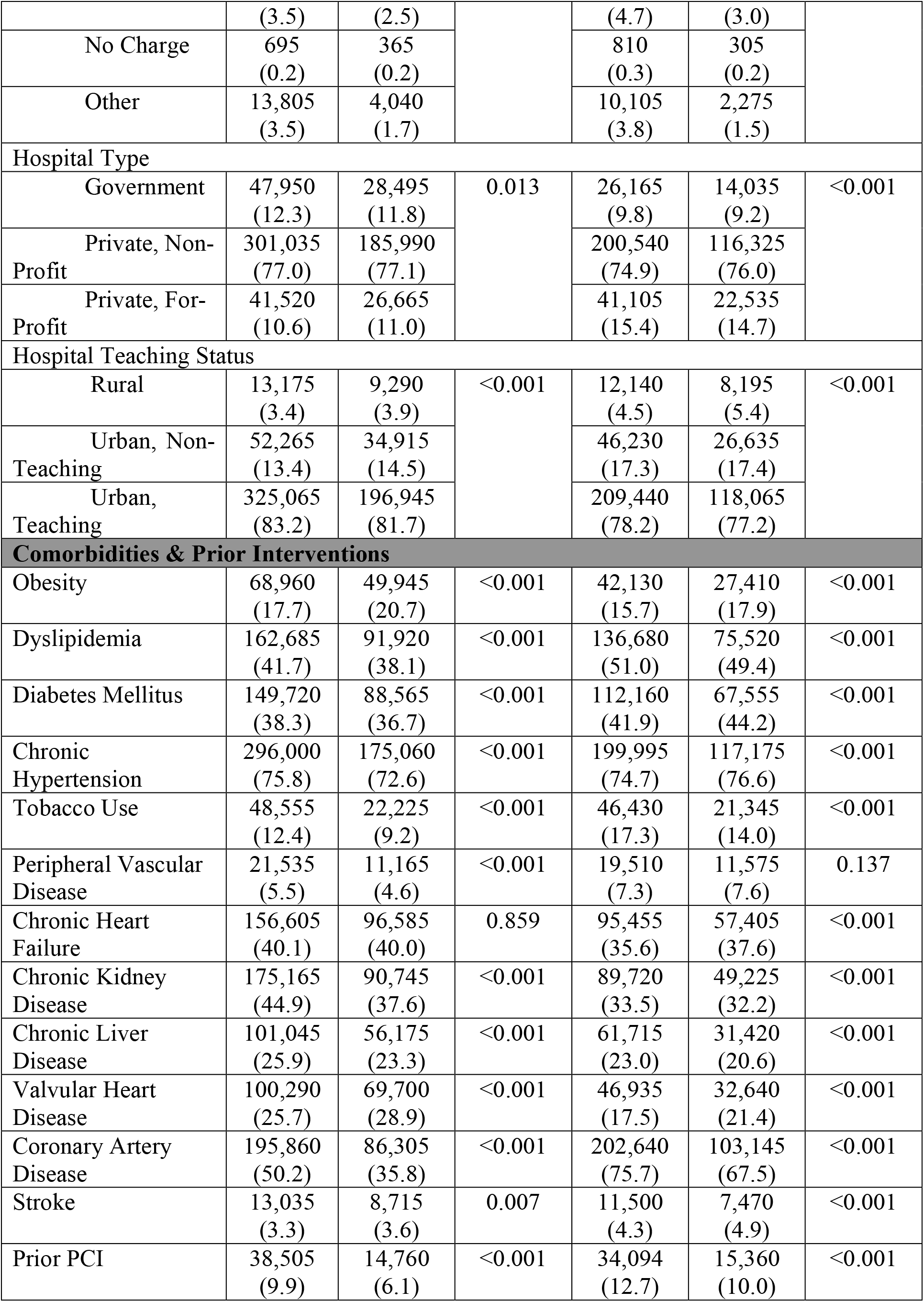
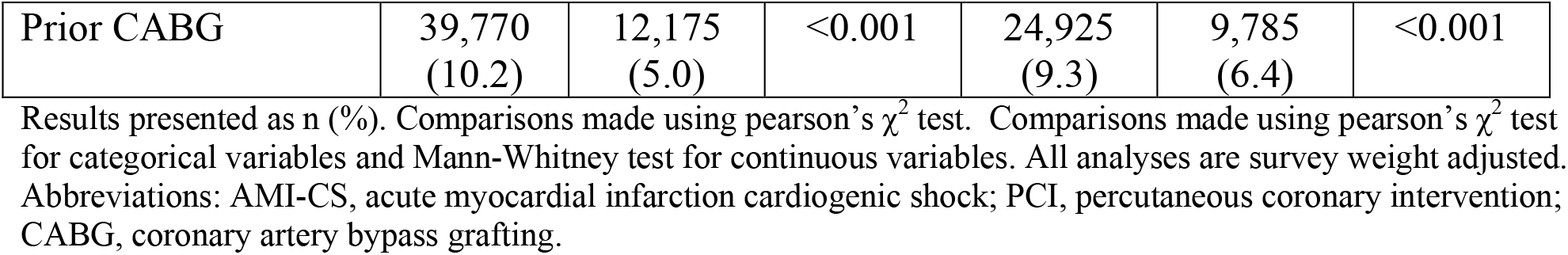
Baseline characteristics of patients stratified by CS type and sex (weighted).

### Procedural and Clinical Outcomes

For both types of CS, women had significantly lower frequencies of almost every type of intervention compared to men (p<0.001), the only exception being heart transplant for AMI-CS (**Table 2**). Women had higher frequencies of in-hospital mortality, use of mechanical ventilation, and stroke during hospitalization, and lower frequencies of major bleeding events and acute kidney injury compared to men. Hospital length of stay and total charges were significantly lower for women than men. These trends were observed after adjusting for baseline characteristics and comorbidities as well (**Tables S2-3**).

**Table 2.**
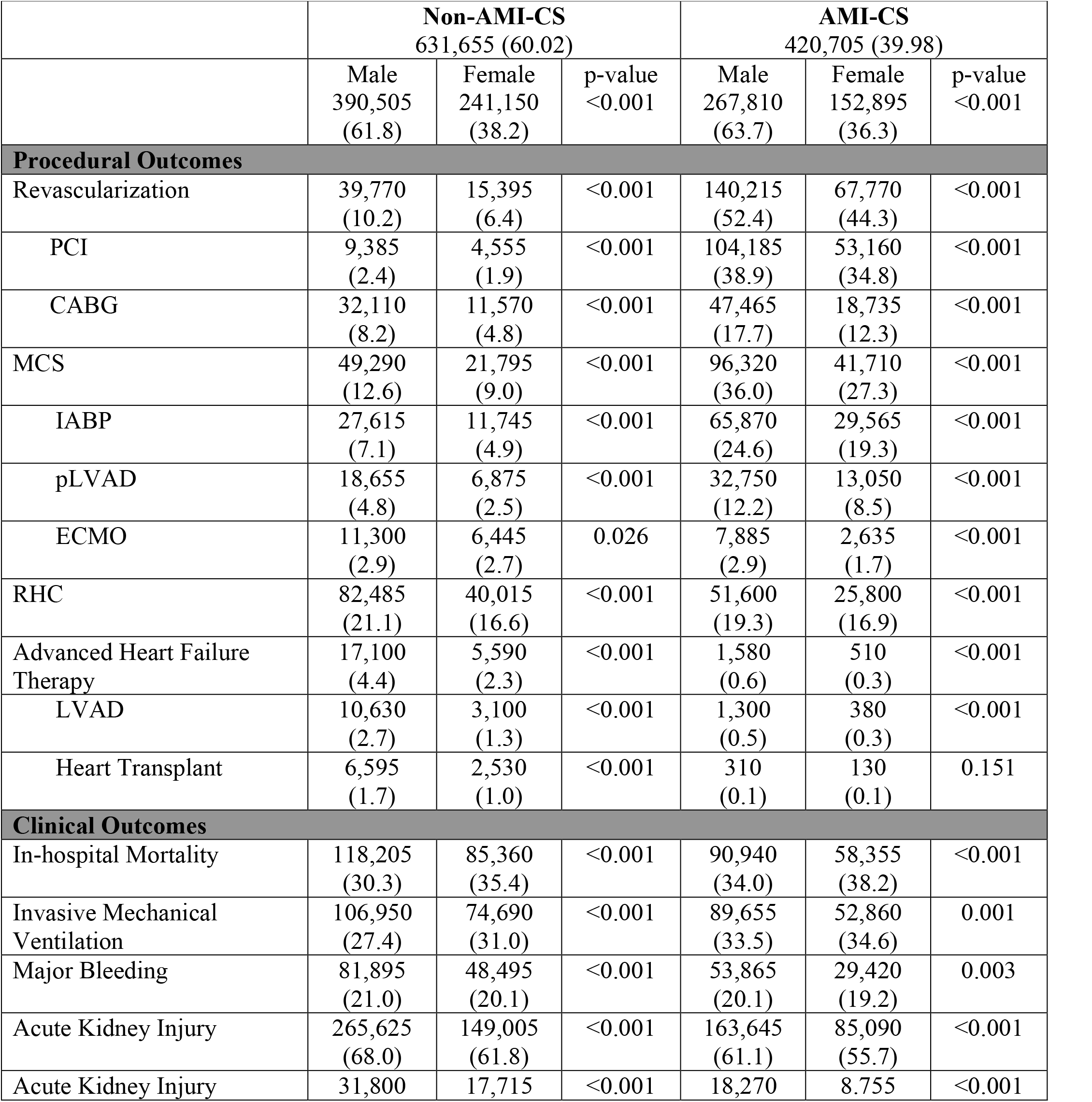
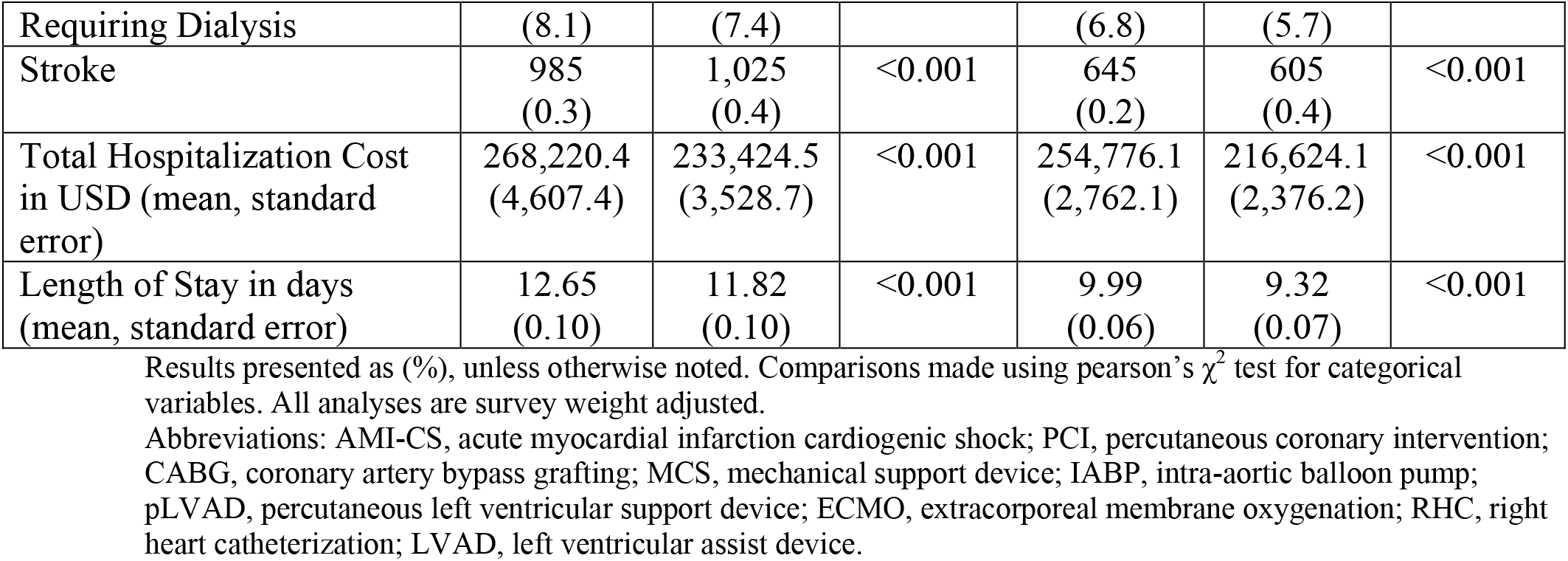
Frequency of procedural and clinical outcomes stratified by sex and CS type.

### Outcomes after Intervention for AMI-CS

Women with AMI-CS undergoing revascularization had higher mortality (aOR: 1.15; 95% CI: 1.09-1.22; p<0.001), were more likely to require mechanical ventilation (aOR: 1.07; 95% CI: 1.02-1.12; p=0.003), and have a stroke (aOR: 1.62; 95% CI: 1.12-2.35; p<0.001) during their hospitalization compared to men (**Table 3**). When MCS was utilized, women were more likely to have higher mortality (adjusted odds ratio (aOR): 1.15; 95% CI: 1.08-1.22; p<0.001), be placed on mechanical ventilation (aOR: 1.12; 95% CI: 1.06-1.19; p<0.001), or have a major bleeding event (aOR: 1.08; 95% CI: 1.01-1.15; p=0.01) during their hospitalization than men. After RHC, women were more likely to die (aOR: 1.10; 95% CI: 1.02-1.19; p<0.001) or be placed on mechanical ventilation (aOR: 1.12; 95% CI: 1.04-1.21; p=0.002) during their hospitalization than men. No significant differences in adverse clinical outcomes between men and women receiving advanced heart failure therapy were noted.

**Table 3.** Odds ratios of clinical outcomes in women versus men with AMI-CS based on first intervention type received.

| Outcomes in Women compared to Men | Unadjusted Odds Ratio | p-value | Model 1 (Only Demographics) Odds Ratio | p-value | Model 2 (All Baseline Characteristics) Odds Ratio | p-value |
| --- | --- | --- | --- | --- | --- | --- |
| **Revascularization** |  |  |  |  |  |  |
| In-hospital Mortality | 1.23 (1.18-1.29) | <0.001 | 1.13 (1.08-1.19) | <0.001 | 1.15 (1.09-1.22) | <0.001 |
| IMV | 1.06 (1.01-1.10) | 0.011 | 1.06 (1.01-1.10) | 0.013 | 1.07 (1.02-1.12) | 0.003 |
| Major Bleeding | 1.02 (0.97-1.07) | 0.322 | 1.02 (0.98-1.08) | 0.245 | 0.98 (0.94-1.04) | 0.689 |
| AKI | 0.79 (0.76-0.83) | <0.001 | 0.73 (0.70-0.76) | <0.001 | 0.71 (0.67-0.74) | <0.001 |
| AKI Requiring Dialysis | 0.89 (0.81-0.98) | 0.021 | 0.87 (0.79-0.97) | 0.013 | 0.86 (0.77-0.96) | 0.007 |
| Stroke | 1.78 (1.25-3.52) | 0.001 | 1.67 (1.16-2.40) | 0.006 | 1.62 (1.12-2.35) | 0.010 |
| **MCS** |  |  |  |  |  |  |
| In-hospital Mortality | 1.19 (1.13-1.26) | <0.001 | 1.12 (1.06-1.19) | <0.001 | 1.15 (1.08-1.22) | <0.001 |
| IMV | 1.11 (1.05-1.17) | <0.001 | 1.10 (1.04-1.17) | 0.001 | 1.12 (1.06-1.19) | <0.001 |
| Major Bleeding | 1.10 (1.04-1.17) | 0.001 | 1.11 (1.04-1.18) | 0.001 | 1.08 (1.01-1.15) | 0.015 |
| AKI | 0.78 (0.74-0.82) | <0.001 | 0.74 (0.70-0.78) | <0.001 | 0.72 (0.68-0.76) | <0.001 |
| AKI Requiring Dialysis | 0.85 (0.77-0.94) | 0.003 | 0.84 (0.76-0.94) | 0.002 | 0.83 (0.74-0.93) | 0.002 |
| Stroke | 1.52 (0.92-2.50) | 0.104 | 1.46 (0.87-2.45) | 0.153 | 1.33 (0.77-2.29) | 0.309 |
| **RHC** |  |  |  |  |  |  |
| In-hospital Mortality | 1.12 (1.04-1.20) | 0.001 | 1.06 (0.99-1.15) | 0.085 | 1.10 (1.02-1.19) | 0.008 |
| IMV | 1.09 (1.02-1.18) | 0.008 | 1.11 (1.04-1.20) | 0.003 | 1.12 (1.04-1.21) | 0.002 |
| Major Bleeding | 1.03 (0.96-1.11) | 0.357 | 1.05 (0.98-1.14) | 0.158 | 1.03 (0.95-1.12) | 0.378 |
| AKI | 0.71 (0.66-0.76) | <0.001 | 0.68 (0.63-0.73) | <0.001 | 0.69 (0.64-0.75) | <0.001 |
| AKI Requiring Dialysis | 0.76 (0.67-0.86) | <0.001 | 0.73 (0.64-0.83) | <0.001 | 0.76 (0.66-0.87) | <0.001 |
| Stroke | 1.12 (0.63-1.99) | 0.687 | 1.06 (0.59-1.90) | 0.837 | 0.96 (0.52-1.77) | 0.901 |
| **Advanced Heart Failure Therapy** |  |  |  |  |  |  |
| In-hospital Mortality | 1.13 (0.58-2.20) | 0.705 | 1.12 (0.48-2.59) | 0.790 | 1.37 (0.54-3.50) | 0.503 |
| IMV | 1.17 (0.63-2.15) | 0.618 | 1.11 (0.55-2.25) | 0.751 | 1.16 (0.58-2.34) | 0.667 |
| Major Bleeding | 1.26 (0.77-2.08) | 0.352 | 1.64 (0.91-2.96) | 0.096 | 1.59 (0.87-2.89) | 0.126 |
| AKI | 0.77 (0.44-1.35) | 0.363 | 0.85 (0.48-1.51) | 0.579 | 0.81 (0.41-1.58) | 0.525 |
| AKI Requiring Dialysis | 1.21 (0.57-2.58) | 0.616 | 1.53 (0.61-3.87) | 0.360 | 1.76 (0.58-5.33) | 0.317 |
| Stroke | 1.55 (0.14-17.6) | 0.721 | 9.66 (0.52-179) | 0.124 | N/A | N/A |

All analyses involve multivariate logistic regression and are survey weight adjusted. Model 1 includes adjustment with year of NIS data, age group, race, household income quartile, insurance status, hospital type and teaching status. Model 2 includes adjustment with all model 1 covariates as well as past medical history of diabetes mellitus, hypertension, dyslipidemia, tobacco use, obesity, chronic kidney disease, liver disease, peripheral vascular disease, coronary artery disease, chronic heart failure, valvular heart disease, stroke, prior PCI, and prior CABG. Abbreviations: AMI-CS, acute myocardial infarction cardiogenic shock; MCS, mechanical support device; RHC, right heart catheterization; IMV, invasive mechanical ventilation; AKI, acute kidney injury.

### Outcomes after Intervention for non-AMI-CS

Following revascularization, women with non-AMI-CS were more likely to die (aOR: 1.38; 95% CI: 1.23-1.56; p<0.001) and be placed on mechanical ventilation (aOR: 1.11; 95% CI: 1.00-1.24; p=0.050) during their hospitalization than men (**Table 4**). Women receiving MCS were more likely to die (aOR: 1.31; 95% CI: 1.21-1.43; p<0.001), be placed on mechanical ventilation (aOR: 1.28; 95% CI: 1.18-1.40; p<0.001), or have a major bleeding event (aOR: 1.10; 95% CI: 1.02-1.18; p=0.012) during their hospitalization than men. The same was true after RHC for in-hospital mortality (aOR: 1.32; 95% CI: 1.22-1.43; p<0.001) and mechanical ventilation usage (aOR: 1.27; 95% CI: 1.17-1.36; p<0.001). No significant differences in adverse clinical outcomes between men and women receiving advanced heart failure therapy were noted.

**Table 4.** Odds ratios of clinical outcomes in women versus men with Non-AMI-CS based on first intervention type received.

| Outcomes in Women compared to Men | Unadjusted Odds Ratio | p-value | Model 1 (Only Demographics) Odds Ratio | p-value | Model 2 (All Baseline Characteristics) Odds Ratio | p-value |
| --- | --- | --- | --- | --- | --- | --- |
| **Revascularization** |  |  |  |  |  |  |
| In-hospital Mortality | 1.43 (1.28-1.59) | <0.001 | 1.37 (1.22-1.54) | <0.001 | 1.38 (1.23-1.56) | <0.001 |
| IMV | 1.11 (1.00-1.23) | 0.033 | 1.10 (0.99-1.23) | 0.060 | 1.11 (1.00-1.24) | 0.050 |
| Major Bleeding | 1.11 (1.02-1.21) | 0.011 | 1.16 (1.06-1.27) | 0.001 | 1.09 (0.99-1.19) | 0.068 |
| AKI | 0.86 (0.79-0.94) | 0.001 | 0.86 (0.78-0.94) | 0.001 | 0.82 (0.74-0.90) | <0.001 |
| AKI Requiring Dialysis | 1.01 (0.85-1.21) | 0.856 | 0.99 (0.82-1.19) | 0.924 | 0.95 (0.78-1.16) | 0.638 |
| Stroke | 1.29 (0.67-2.48) | 0.439 | 1.42 (0.71-2.86) | 0.318 | 1.32 (0.66-2.65) | 0.423 |
| **MCS** |  |  |  |  |  |  |
| In-hospital Mortality | 1.30 (1.20-1.40) | <0.001 | 1.30 (1.20-1.42) | <0.001 | 1.31 (1.21-1.43) | <0.001 |
| IMV | 1.27 (1.17-1.37) | <0.001 | 1.27 (1.17-1.39) | <0.001 | 1.28 (1.18-1.40) | <0.001 |
| Major Bleeding | 1.12 (1.05-1.20) | 0.001 | 1.14 (1.06-1.23) | <0.001 | 1.10 (1.02-1.18) | 0.012 |
| AKI | 0.68 (0.63-0.73) | <0.001 | 0.67 (0.62-0.73) | <0.001 | 0.66 (0.60-0.73) | <0.001 |
| AKI Requiring Dialysis | 0.91 (0.82-1.02) | 0.114 | 0.91 (0.81-1.02) | 0.126 | 0.94 (0.82-1.06) | 0.324 |
| Stroke | 1.64 (0.96-2.83) | 0.070 | 1.69 (0.96-2.98) | 0.068 | 1.64 (0.93-2.89) | 0.089 |
| **RHC** |  |  |  |  |  |  |
| In-hospital Mortality | 1.30 (1.21-1.39) | <0.001 | 1.31 (1.22-1.41) | <0.001 | 1.32 (1.22-1.43) | <0.001 |
| IMV | 1.30 (1.21-1.39) | <0.001 | 1.31 (1.22-1.41) | <0.001 | 1.27 (1.17-1.36) | <0.001 |
| Major Bleeding | 1.01 (0.95-1.08) | 0.609 | 1.02 (0.96-1.09) | 0.427 | 0.98 (0.91-1.04) | 0.507 |
| AKI | 0.69 (0.65-0.73) | <0.001 | 0.68 (0.64-0.72) | <0.001 | 0.73 (0.68-0.77) | <0.001 |
| AKI Requiring Dialysis | 0.95 (0.86-1.04) | 0.285 | 0.95 (0.85-1.04) | 0.273 | 1.00 (0.90-1.11) | 0.970 |
| Stroke | 1.45 (0.89-2.37) | 0.127 | 1.54 (0.93-2.56) | 0.090 | 1.55 (0.91-2.63) | 0.104 |
| **Advanced Heart Failure Therapy** |  |  |  |  |  |  |
| In-hospital Mortality | 1.05 (0.81-1.36) | 0.671 | 1.13 (0.86-1.50) | 0.361 | 1.20 (0.88-1.63) | 0.242 |
| IMV | 1.00 (0.81-1.23) | 0.952 | 0.99 (0.79-1.24) | 0.969 | 1.05 (0.84-1.31) | 0.635 |
| Major Bleeding | 1.03 (0.88-1.20) | 0.665 | 0.99 (0.84-1.16) | 0.906 | 1.02 (0.87-1.20) | 0.768 |
| AKI | 0.62 (0.52-0.72) | <0.001 | 0.61 (0.52-0.71) | <0.001 | 0.68 (0.57-0.81) | <0.001 |
| AKI Requiring Dialysis | 0.79 (0.59-1.05) | 0.115 | 0.76 (0.57-1.03) | 0.081 | 0.79 (0.57-1.09) | 0.157 |
| Stroke | 1.45 (0.65-3.20) | 0.354 | 1.39 (0.60-3.22) | 0.432 | 1.44 (0.62-3.35) | 0.393 |

All analyses involve multivariate logistic regression and are survey weight adjusted. Model 1 includes adjustment with year of NIS data, age group, race, household income quartile, insurance status, hospital type and teaching

### Incidence and Mortality Following Early Intervention

For both types of shock, women had a lower frequency of receiving CABG, IABP, pLVAD, and RHC within 24 hours of admission compared to men (**Figure 1**). Women with AMI-CS additionally had lower frequencies of undergoing placement on ECMO and PCI within 24 hours. This was similarly reflected after adjusting for covariates, with women with AMI-CS being less likely to receive any type of revascularization, MCS, or RHC than men, and women with non-AMI-CS being less likely to receive any type of revascularization, IABP, pLVAD, RHC, or LVAD than men (**Figure 2**). In-hospital mortality did not change significantly between men and women with AMI-CS undergoing early revascularization, pLVAD, ECMO, or RHC (**Figure 3**). For those with non-AMI-CS, in-hospital mortality did not change significantly when treated early with PCI, pLVAD, ECMO, or RHC (**Figure 3**). For AMI-CS, women receiving early interventions of any revascularization or within 24 hours were far less likely to die during their hospitalization than women who received intervention after 24 hours or not at all; meanwhile, for non-AMI-CS, this was the case for women receiving early CABG, IABP, or RHC (**Table 5**).

**Table 5.** Odds ratios of in-hospital mortality in women receiving early (within 24 hours) versus late or no intervention by CS type.

| In-hospital Mortality in Women Receiving Early Intervention compared to Women Not Receiving Early Intervention | Unadjusted Odds Ratio | p-value | Model 1 (Only Demographics) Odds Ratio | p-value | Model 2 (All Baseline Characteristics) Odds Ratio | p-value |
| --- | --- | --- | --- | --- | --- | --- |
| **AMI-CS** |  |  |  |  |  |  |
| Revascularization | 0.65 (0.61-0.69) | <0.001 | 0.66 (0.62-0.70) | <0.001 | 0.77 (0.72-0.82) | <0.001 |
| PCI | 0.66 (0.63-0.71) | <0.001 | 0.68 (0.64-0.72) | <0.001 | 0.79 (0.74-0.85) | <0.001 |
| CABG | 0.63 (0.55-0.73) | <0.001 | 0.63 (0.55-0.74) | <0.001 | 0.69 (0.59-0.82) | <0.001 |
| MCS | 0.87 (0.82-0.93) | <0.001 | 0.94 (0.88-1.00) | 0.068 | 1.06 (0.99-1.14) | 0.073 |
| IABP | 0.69 (0.64-0.75) | <0.001 | 0.72 (0.67-0.78) | <0.001 | 0.83 (0.77-0.91) | <0.001 |
| pLVAD | 1.50 (1.35-1.68) | <0.001 | 1.67 (1.49-1.88) | <0.001 | 1.78 (1.58-2.01) | <0.001 |
| ECMO | 1.71 (1.32-2.2) | <0.001 | 2.38 (1.83-3.10) | <0.001 | 2.15 (1.62-2.85) | <0.001 |
| RHC | 0.87 (0.79-0.96) | 0.005 | 0.90 (0.819-0.99) | 0.048 | 0.979 (0.88-1.08) | 0.688 |
| **Non-AMI-CS** |  |  |  |  |  |  |
| Revascularization | 0.42 (0.37-0.49) | <0.001 | 0.42 (0.36-0.49) | <0.001 | 0.57 (0.49-0.67) | <0.001 |
| PCI | 0.76 (0.60-0.95) | 0.017 | 0.70 (0.55-0.88) | 0.003 | 0.87 (0.68-1.12) | 0.296 |
| CABG | 0.32 (0.27-0.39) | <0.001 | 0.33 (0.28-0.40) | <0.001 | 0.46 (0.38-0.56) | <0.001 |
| MCS | 0.92 (0.82-1.01) | 0.109 | 0.97 (0.87-1.09) | 0.663 | 1.05 (0.94-1.18) | 0.360 |
| IABP | 0.64 (0.54-0.74) | <0.001 | 0.65 (0.55-0.77) | <0.001 | 0.78 (0.67-0.93) | 0.005 |
| pLVAD | 1.41 (1.14-1.74) | 0.001 | 1.48 (1.19-1.84) | <0.001 | 1.56 (1.24-1.96) | <0.001 |
| ECMO | 1.31 (1.10-1.57) | 0.002 | 1.52 (1.26-1.82) | <0.001 | 1.40 (1.15-1.69) | 0.001 |
| RHC | 0.42 (0.37-0.46) | <0.001 | 0.45 (0.39-0.50) | <0.001 | 0.50 (0.44-0.57) | <0.001 |

**Figure 1.**
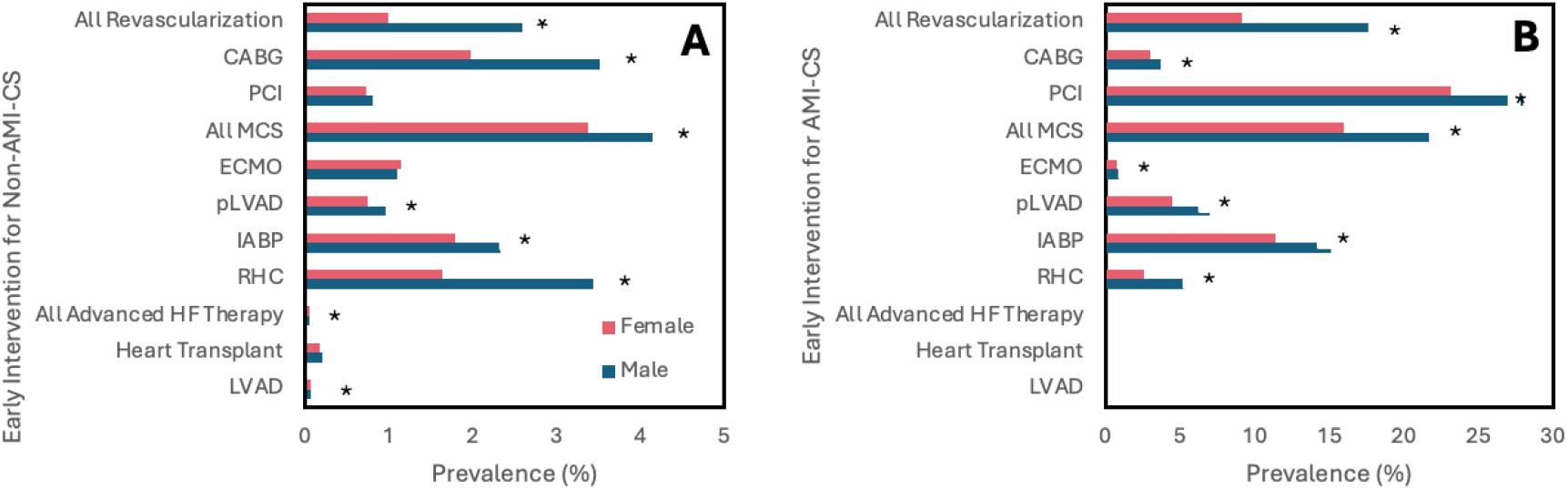
Frequency of early intervention within 24 hours stratified by sex and CS type. Unadjusted results. Figure 1A and 1B shows results for non-AMI-CS and AMI-CS, respectively. Abbreviations: AMI-CS, acute myocardial infarction cardiogenic shock; PCI, percutaneous coronary intervention; CABG, coronary artery bypass grafting; MCS, mechanical support device; IABP, intra-aortic balloon pump; pLVAD, percutaneous left ventricular support device; ECMO, extracorporeal membrane oxygenation; RHC, right heart catheterization; LVAD, left ventricular assist device.*p<0.001

**Figure 2.**
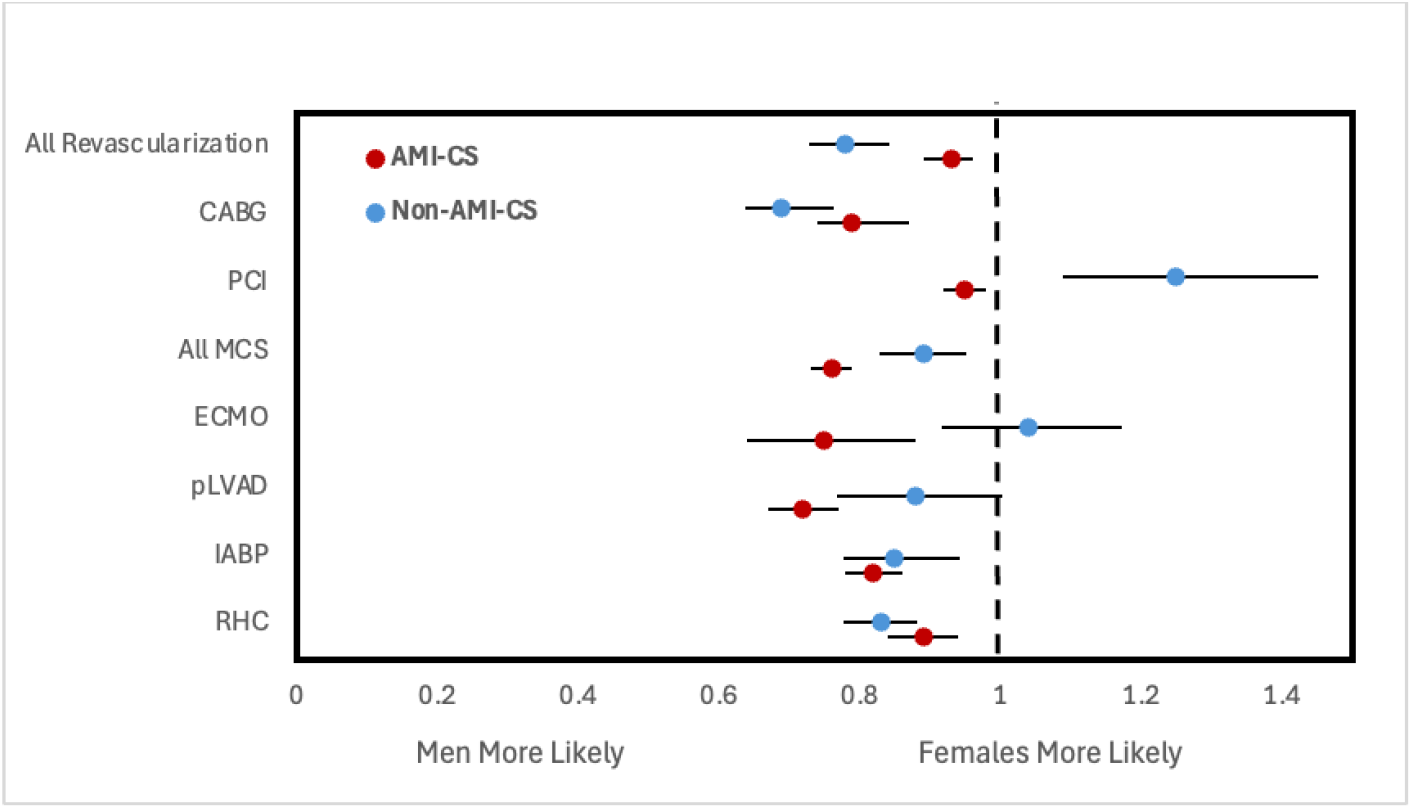
Adjusted odds ratios of receiving early intervention within 24 hours of admission stratified by CS type.All analyses involve multivariate logistic regression and are survey weight adjusted. Odds ratios reported are adjusted for year of NIS data, age group, race, household income quartile, insurance status, hospital type and teaching status as well as past medical history of diabetes mellitus, hypertension, dyslipidemia, tobacco use, obesity, chronic kidney disease, liver disease, peripheral vascular disease, coronary artery disease, chronic heart failure, valvular heart disease, stroke, prior PCI, and prior CABG.Note: advanced heart failure therapies data not reported due to low counts preventing proper construction of logistic regression. Abbreviations: AMI-CS, acute myocardial infarction cardiogenic shock; PCI, percutaneous coronary intervention; CABG, coronary artery bypass grafting; MCS, mechanical support device; IABP, intra-aortic balloon pump; pLVAD, percutaneous left ventricular support device; ECMO, extracorporeal membrane oxygenation; RHC, right heart catheterization.

**Figure 3.**
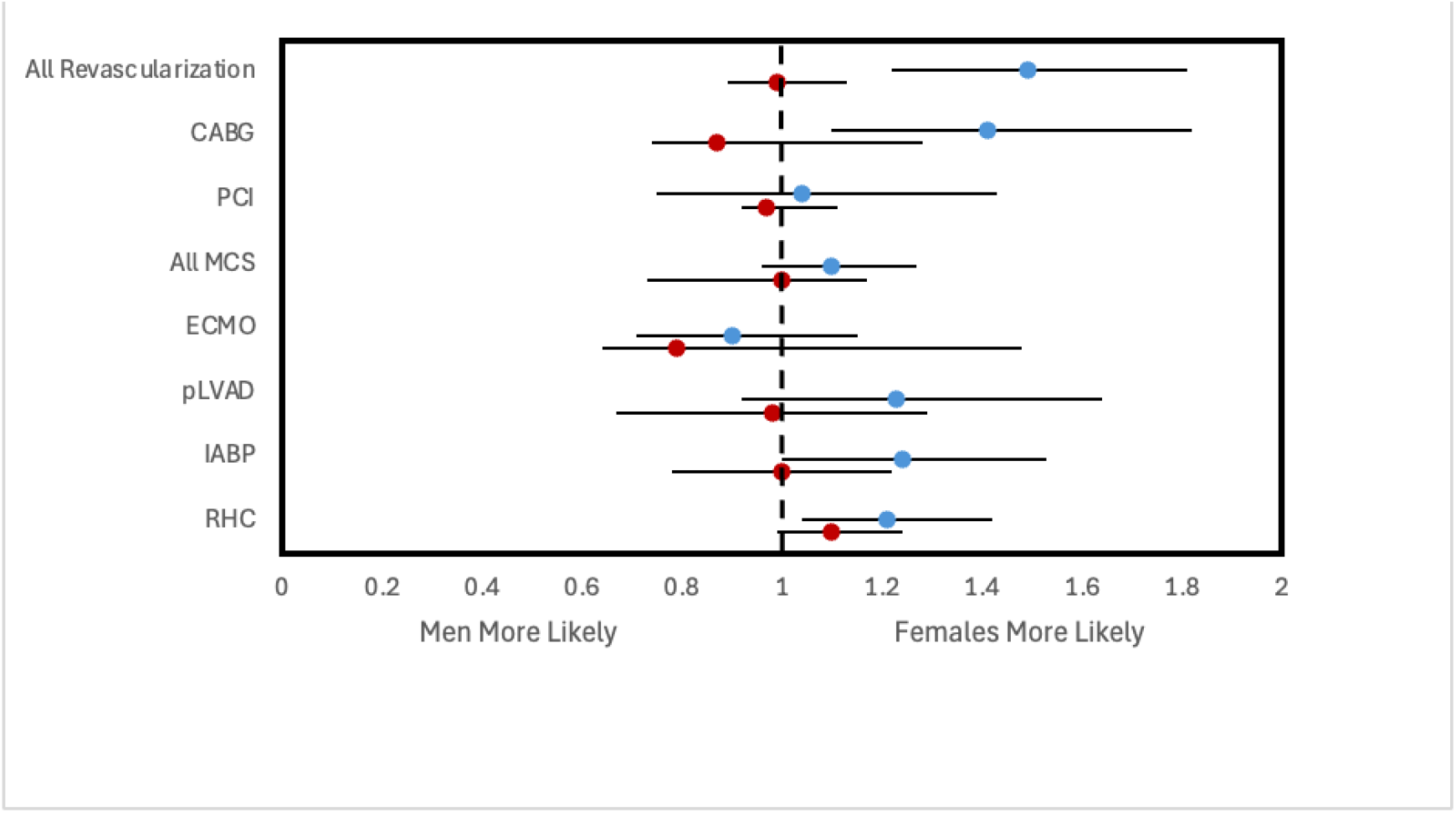
Adjusted odds ratios of in-hospital mortality following early intervention within 24 hours of admission stratified by CS type.All analyses involve multivariate logistic regression and are survey weight adjusted. Each regression is performed on a dataset filtered to only include CS patients who received a particular intervention within 24 hours. Odds ratios reported are adjusted for year of NIS data, age group, race, household income quartile, insurance status, hospital type and teaching status as well as past medical history of diabetes mellitus, hypertension, dyslipidemia, tobacco use, obesity, chronic kidney disease, liver disease, peripheral vascular disease, coronary artery disease, chronic heart failure, valvular heart disease, stroke, prior PCI, and prior CABG.Note: advanced heart failure therapies data not reported due to low counts preventing proper construction of logistic regression. Abbreviations: AMI-CS, acute myocardial infarction cardiogenic shock; PCI, percutaneous coronary intervention; CABG, coronary artery bypass grafting; MCS, mechanical support device; IABP, intra-aortic balloon pump; pLVAD, percutaneous left ventricular support device; ECMO, extracorporeal membrane oxygenation; RHC, right heart catheterization.

## Discussion

To our knowledge, this study is the first to examine sex differences in management and outcomes of patients with AMI-CS and non-AMI-CS utilizing time to intervention. We found that (1) women with either type of CS are less likely to undergo interventions during hospitalization than men; (2) even after receiving intervention, in-hospital mortality, mechanical ventilation, and major bleeding are more likely to occur in women with CS than men; (3) among patients undergoing intervention, women are less likely to have an intervention performed within 24 hours of admission; (4) and with early intervention, there was no difference in mortality among women and men.

Previous studies have demonstrated that women are less likely to receive MCS therapies in CS and experience a 10% higher mortality risk.^11^ Not only did we confirm these findings, but we also found that women had significantly lower frequencies of nearly all types of interventions compared to men and were less likely to receive these interventions early. This was associated with higher frequencies of in-hospital mortality, invasive mechanical ventilation, and stroke during hospitalization. Prior analyses of NIS data from 2006 to 2015 found that, for AMI-CS, women were more likely to die during their hospitalization than men (aOR 1.11, 95% CI: 1.06-1.16 p < 0.001).^15^ Despite significant recognition of sex disparities in management and mortality rates of many conditions, our analysis shows that the situation has yet to improve significantly in the subsequent decade for CS.

The results of this study suggest that ongoing sex disparities in mortality and outcomes of CS may be related to time to intervention. When women and men are similarly treated with early intervention for CS, outcomes are comparable. Additionally, women with CS undergoing any revascularization, IABP, or RHC within 24 hours of admission were far less likely to die during their hospitalization than women who received intervention after 24 hours or not at all. Both of these findings point to late intervention as a plausible explanation for the observed outcome differences and demonstrate the tangible impact of timely intervention in narrowing the sex gap in clinical outcomes for CS.

Research has extensively explored the elevated mortality in women with CS, especially within the context of AMI. One hypothesis posits that women in low-output states are less likely to be recognized by providers as experiencing CS. This is supported by studies on sex-based differences in time to treatment for AMI, showing delays at every stage of care (from symptom onset to EMS call, EMS to hospital admission, and hospital admission to reperfusion therapy).^16^ Early intervention has been shown to be crucial in the treatment of CS, lending to better post-procedure outcomes and lower rates of in- and out-hospital mortality.^17,18^ Reasons for this include improved perfusion resulting in the prevention of irreversible ischemic damage and pathologic cardiac remodeling.

Clinically, improving recognition of low-output states in women can lead to decreased time to treatment for CS. When looking at the temporal trends of women undergoing AMI, female sex has been associated with a greater time to reperfusion even when adjusting for all baseline characteristics and types of reperfusion therapy used.^19,20^ It is equally important to note that when performing a clinical assessment of suspected AMI, symptom presentation varies not only by sex, but also by race; for example, Black women are more likely to present with GI distress, while White women are more likely to present with typical chest pain.^21^ Noting these differences in symptom presentation is significant, as an atypical presentation with acute-onset GI distress can modify the differential to include a diagnosis of GERD or peptic ulcer disease, which would subsequently contribute to poorer outcomes, especially in Black women.

### Limitations

There were a few limitations that were present in this study. The diagnosis of CS was based on ICD-CM codes used for billing purposes, which are susceptible to erroneous coding. However, previous studies comparing ICD diagnosis of CS to physician adjudication in hospital systems that validate these codes have demonstrated that ICD-CM codes are reliably coded and can accurately predict mortality in the context of MI and CS.^22,23^ Also, the NIS database recorded time in units of days as opposed to hours, so more exact timing information was not available. Data included information on hospitalizations as opposed to patient-level data, which opens the possibility of repeat patients in the sample. Additionally, specific data such as vital signs, laboratory, and echocardiogram data were not available. Finally, there was a lack of information on post-discharge outcomes, which could have provided further insight into post-intervention results from the management of CS.

## Conclusions

Our study found that, despite stark differences in access to early intervention for CS between men and women, in-hospital mortality did not differ significantly between sexes when treated equally within 24 hours of admission. Early intervention plays a role in mitigating sex-based differences in CS outcomes and should be prioritized in the management of CS. Future research should focus on recognizing the atypical presentations of CS in women, identifying signs and symptoms earlier in the disease course, and validating our findings in other cohorts.

## Data Availability

The data used in this study was adapted from the National Inpatient Sample database and is, therefore, unavailable for public posting. The data is available for purchase on the Agency for Healthcare Research and Quality website.

https://hcup-us.ahrq.gov/db/nation/nis/nisdbdocumentation.jsp

## Abbreviations

CS: Cardiogenic Shock
AKI: Acute Kidney Injury
AMI: Acute Myocardial Infarction
MCS: Mechanical Circulatory Support
RHC: Right Heart Catheterization
HCUP: Healthcare Cost and Utilization Project
NIS: National Inpatient Sample
PCI: Percutaneous Intervention
CABG: Coronary Artery Bypass Graft
IABP: Intra-Aortic Balloon Pump
pLVAD: Percutaneous Left Ventricular Assist Device
LVAD: Left Ventricular Assist Device
ECMO: Extracorporeal Membrane Oxygenation
aOR: Adjusted Odds Ratio

## Acknowledgements

None

